# Non-patient related SARS-CoV-2 exposure to colleagues and household members impose the highest infection risk for hospital employees with and without patient contact in a German university hospital: follow-up of the prospective Co-HCW Seroprevalence study

**DOI:** 10.1101/2022.10.05.22280728

**Authors:** Christina Bahrs, Sebastian Weis, Miriam Kesselmeier, Juliane Ankert, Stefan Hagel, Stephanie Beier, Jens Maschmann, Andreas Stallmach, Andrea Steiner, Michael Bauer, Wilhelm Behringer, Michael Baier, Cora Richert, Florian Zepf, Martin Walter, André Scherag, Michael Kiehntopf, Bettina Löffler, Mathias W. Pletz

**Author notes:** Corresponding author: Christina Bahrs, MD, Institute of Infectious Diseases and Infection Control, Jena University Hospital/Friedrich-Schiller-University, Am Klinikum 1, 07747 Jena, Germany.

## Abstract

**Background:** The Co-HCW study is a prospective, longitudinal single center observational study on the SARS-CoV-2 seroprevalence and infection status in staff members of Jena University Hospital (JUH) in Jena, Germany.

**Material and Methods:** This follow-up study covers the observation period from 19^th^ May 2020 to 22^nd^ June 2021. At each out of three voluntary study visits, participants filled out a questionnaire on individual SARS-CoV-2 exposure. In addition, serum samples to assess specific SARS-CoV-2 antibodies were collected. Participants with antibodies against nucleocapsid and/or spike protein without previous vaccination and/or a reported positive SARS-CoV-2 PCR test were regarded as participants with detected SARS-CoV-2 infection. Multivariable logistic regression modeling was applied to identify potential risk factors for infected compared to non-infected participants.

**Results:** Out of 660 participants that were included during the first study visit, 406 participants (61.5%) were eligible for final analysis as they did not change the COVID-19 risk area (high-risk n=76; intermediate-risk n=198; low-risk n=132) during the study. Forty-four participants (10.8%, 95% confidence interval (95%CI) 8.0%-14.3%) had evidence of a current or past SARS-CoV-2 infection detected by serology (n=40) and/or PCR (n=28). No association of any SARS-CoV-2 infection with the COVID-19 risk group according to working place could be detected. But exposure to a SARS-CoV-2 positive household member (adjusted OR (AOR) 4.46, 95%CI 2.06-9.65) or colleague (AOR 2.30, 95%CI 1.10-4.79) significantly increased the risk of a SARS-CoV-2 infection.

**Conclusion:** Our results demonstrate that non-patient-related SARS-CoV-2 exposure imposed the highest infection risk in hospital staff members of JUH.

## Introduction

Healthcare workers (HCW) across the world are at high risk to acquire coronavirus disease 2019 (COVID-19) caused by severe acute respiratory syndrome coronavirus 2 (SARS-CoV-2) ^1-3^, as they are directly or indirectly exposed to infectious material ^3^ while caring for patients suffering from the coronavirus disease 2019 (COVID-19) ^4^. Transmission of SARS-CoV-2 occurs primarily via inhalation of, or inoculation with infectious small liquid particles ranging from larger respiratory droplets to smaller aerosols in case of close personal contact ^5^. Aerosol transmission in health-care settings may occur in specific situations in which HCW perform medical, aerosol generating procedures but do not use adequate personal protection equipment (PPE) ^5^. With the ongoing COVID-19 pandemic ^6,7^, ensuring the safety of HCW is of utmost relevance ^1,3,5^. Infection control measures, including the use of adequate PPE, hand hygiene, and physical separation are considered essential in reducing nosocomial transmissions ^5,8^. Additionally, vaccination of patients and HCW reduces the risk of acquiring COVID-19 in health care settings.

The city of Jena, with a population of approximately 111,000 inhabitants, hosts the only university hospital of the entire federal state Thuringia (Jena University Hospital, JUH), which is located in central Germany. Besides there is no other hospital in the city of Jena. In March 2020, mandatory masking was implemented for all staff members of JUH, including HCW and administration staff ^9^ aiming to reduce nosocomial SARS-CoV-2 transmissions. Additionally, business trips and participation in presence on conferences or trainings activities outside JUH were prohibited for all employees by the local Medical Executive board. In December 2020, SARS-CoV-2 vaccination was first available and was initially offered to HCW with high risk. Since February 2021, SARS-CoV-2 vaccination was offered to all hospital staff members. The vaccination rate documented by the department of occupational health of JUH in December 2021 was 85% (94% for physicians, 88% for nurses, and 85% for administration staff). We have previously reported a low SARS-CoV-2 point seroprevalence rate of 2.7% among hospital staff (inclusion of first participant: 19^th^ May 2020, inclusion of last participant: 19^th^ June 2020) ^9^, and identified COVID-19 exposure at home as the main risk factor associated with SARS-CoV-2 point seroprevalence. This was prior to availability of SARS-CoV-2 vaccination.

The primary objective of this follow-up study was to assess the SARS-CoV-2 seroprevalence and prevalence of SARS-CoV-2 infection among employees with (HCW) and without patient contact (administration staff) of JUH over a period of 13 months (May 2020 to June 2021). Secondary objectives were to determine individual exposure risk factors, and to compare SARS-CoV-2 infection rates between hospital staff working at different COVID-19 risk areas according to working place.

## Methods

### Study design and setting

The Co-HCW study (SARS-CoV-2 seroprevalence and infection status in hospital staff members at JUH) is a prospective, longitudinal single centre observational cohort study conducted at JUH, a 1,400-bed academic hospital in Germany. The first of three visits (05/2020) has already been published ^9^. This current analysis covers the complete observation period of 11-13 months and includes data from 19^th^ May 2020 to 22^nd^ June 2021. At our hospital, intensive SARS-CoV-2 screening was carried out. Details of the routine PCR screening are described below.

Research was conducted in accordance with the Declaration of Helsinki and national and institutional standards. The study protocol was approved by the local ethics committee of the Friedrich-Schiller-University Jena (approval no. 2020–1774), and the study was registered at the German Clinical Trials Register (DRKS00022432).

### Enrolment and data management

Participants including hospital staff and administration staff were recruited between 19^th^ May 2020 and 19^th^ June 2020. For inclusion and exclusion criteria as well as data management, we refer to the previously published results of the first study visit ^9^. In total, three study visits were offered to all participants. Participation in each study visit was voluntary. The first study visit was performed at inclusion, the second study visit was performed from 6^th^ November 2020 to 26^th^ November 2020, and the third study visit was performed during 26^th^ April 2021 and 22^nd^ June 2021. For the present analysis, only participants were considered who completed the last study visit in 2021 and did not change the COVID-19 risk area according to their risk of a contact with COVID-19 patients at work (low, intermediate and high risk) during the study.

At each study visit, participants had to fill out a questionnaire, and blood samples were collected at the study center, which were then sent to the Department of Clinical Chemistry and Laboratory Medicine of JUH and the Institute of Medical Microbiology of JUH for testing of specific SARS-CoV-2 antibodies by two different immunoassays (see below).

### Questionnaire

As previously described ^9^, the questionnaire included questions on demographics, profession, working area, individual exposure to confirmed COVID-19 cases, return from COVID-19 risk areas, results of previous polymerase chain reaction (PCR) or serology test for COVID-19, clinical symptoms, accidents with biological material and compliance concerning use of PPE in HCW with individual contact with a confirmed COVID-19 patient. Due to the recommendation of the referees of the first peer-review of this study, we additionally included the following parameters in the updated questionnaire for the second and third visit: use of public transport on the way to work, household size, travel to abroad and participation at events with at least five persons. As SARS-CoV-2 vaccination has been available since 27^th^ December 2020, the questionnaire of the last visit was further extended with questions on number and type of SARS-CoV-2 vaccinations.

### PCR Screening

All staff in high-risk areas (intensive care unit, intermediate care unit, emergency department and COVID-19 regular ward) were tested twice a week by PCR. In addition, all staff members were called upon to have a PCR test carried out in case of symptoms of infection and/or after 1 and 5 days of contact with a SARS-CoV-2 infected person at work or at home. Furthermore, in case of nosocomial transmission detected by patient screening, the staff of the respective ward were screened on day 1 and 5.

### SARS-CoV-2 antibody testing

Specific SARS-CoV-2 antibodies in serum samples were detected at each time point deploying the commercially available chemiluminescence-based immunoassay (CLIA) Elecsys Anti-SARS-CoV-2 (Roche, Basel, Switzerland) that uses a recombinant nucleocapsid protein as capture antigen. At the first and second visits the enzyme-linked immunosorbent assay EDI Novel Coronavirus SARS-CoV-2 IgG ELISA (Epitope Diagnostics Inc., San Diego, USA, antigen: recombinant nucleocapsid protein) was performed as a second method. At visit three spike-protein specific IgG antibodies were identified using the CLIA system LIAISON® SARS CoV-2 S1/S2 IgG (DiaSorin, Saluggia, Italy). All serological tests were carried out according to the manufacturers’ instructions. Sensitivities and specificities as provided by the manufacturers are high for all tests (≥97%).

Participants with at least one positive test result for antibodies against nucleocapsid and/or spike protein without previous vaccination and/or a reported positive SARS-CoV-2 PCR test were regarded as participants with detected SARS-CoV-2 infection.

### Outcomes and further definitions

The primary outcome of this follow-up study was to assess the SARS-CoV-2 infection rates using SARS-CoV-2 antibody detecting immunoassays and reported positive SARS-CoV-2 PCR test results. Secondary outcomes were (i) prevalence of SARS-CoV-2 infection in participants stratified by their risk of COVID-19 exposure during work (low, medium and high risk), and (ii) potential risk factors for detected SARS-CoV-2 infection including compliance of HCW in case of an individual reported contact with a confirmed COVID-19 positive patient.

### Statistical analysis

Characteristics of participants are summarized (overall, stratified by test result) as absolute and relative frequencies or as median together with first and third quartile (Q1, Q3). Evidence of any SARS-CoV-2 infection in hospital staff within the observation period is described with absolute and relative frequencies together with 95% Clopper-Pearson confidence intervals (CIs). To compare SARS-CoV-2 infection rates between participants working at different COVID-19 risk areas, and to identify potential risk factors for infected compared to non-infected participants, we apply uni- and multivariable logistic regression modelling with the SARS-CoV-2 infection as dependent variable and the investigated factor as independent variable. In the multivariable models, we adjusted for age and gender. For place of exposure, we considered two additional multivariable models. In the first additional model, we included all places, that were assessed, as independent variables to adjust each investigated place for the respective other places. In the second additional model, we adjusted this model for age and gender. We provide (adjusted) odds ratios (OR) together with 95% CI and p-value.

We applied a two-sided significance level of 0.05 and did not correct for multiple testing as all analyses were considered exploratory. The main analyses were done with R (version 4.0.3), and parts were complemented by SPSS Statistics version 28.0 for Windows (IBM Corp., Armonk, NY, USA).

## Results

### Characteristics of the study population

Out of 660 participants that were analysed during the first study visit, 406 hospital staff members (61.5%) also participated in the third and last study visit and did not change the COVID-19 risk area during the reported 13 months. Of these 406 participants, 91 (22.4%) were males and 315 (77.6%) were females. The median age of the participants was 41.0 (Q1-Q3: 34.0-49.8) years. The most common professions included administration staff (n=132, 32.5%), followed by nurses (n=125, 30.8%), physicians (n=66, 16.3%), reception staff (n=12, 3.0%), nursing assistants (n=10, 2.5%), psychologists (n=10, 2.5%), ergo therapists (n=10, 2.5%), and medical assistants (n=9, 2.2%). Two-hundred twenty-four participants (55.2%) reported direct contact to a confirmed COVID-19 case, whereas 182 participants (44.8%) were not aware of any COVID-19 exposure. Among the 224 staff members with reported COVID-19 exposure, 151 participants (67.4%) had direct contact with a SARS-CoV-2 positive patient, and 60 participants (26.8%) had exposure to a SARS-CoV-2 positive colleague. Direct COVID-19 contact outside the JUH included close contact to a positive household member (n=43, 19.2%), exposure to friends (n=20, 8.9%), exposure during shopping (n=2, 0.9%) and exposure on holiday (n=1, 0.4%). Further details on the participants are provided in Table 1. Any SARS-CoV-2 vaccination prior to the last study visit was reported from 307 participants (75.6%); 177 participants (43.6%) had received two vaccinations (homologous vaccination with a COVID-19 messenger RNA (mRNA) vaccine: n=160; homologous vaccination with the vector-based vaccine ChAdOx1-S: n=7; heterologous vaccination with the vector-based vaccine followed by a mRNA vaccine: n=10) and 130 participants (32.0%) had received one vaccination (COVID-19 mRNA vaccine: n=16; COVID-19 vector-based vaccine ChAdOx1-S: n=114).

**Table 1.**
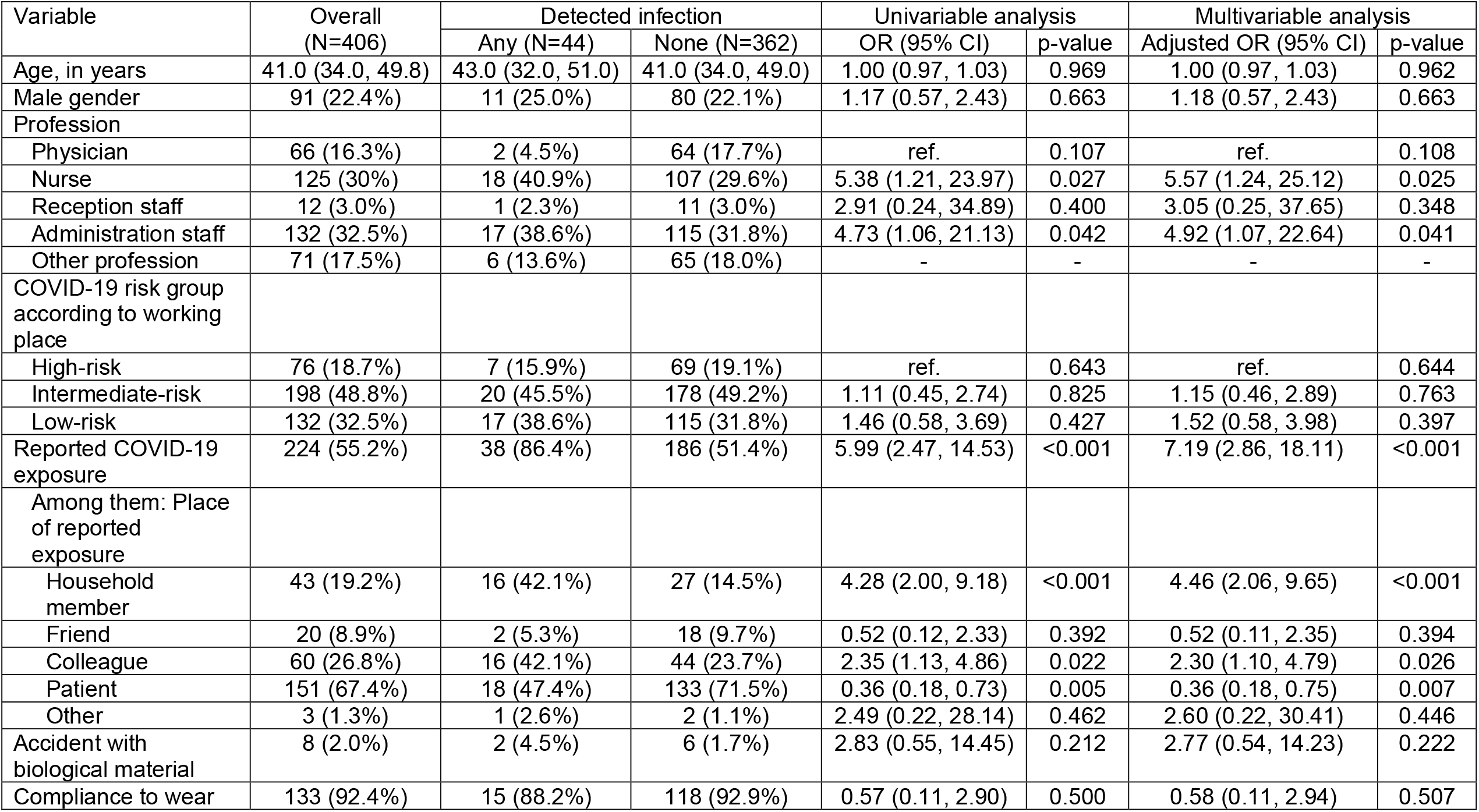
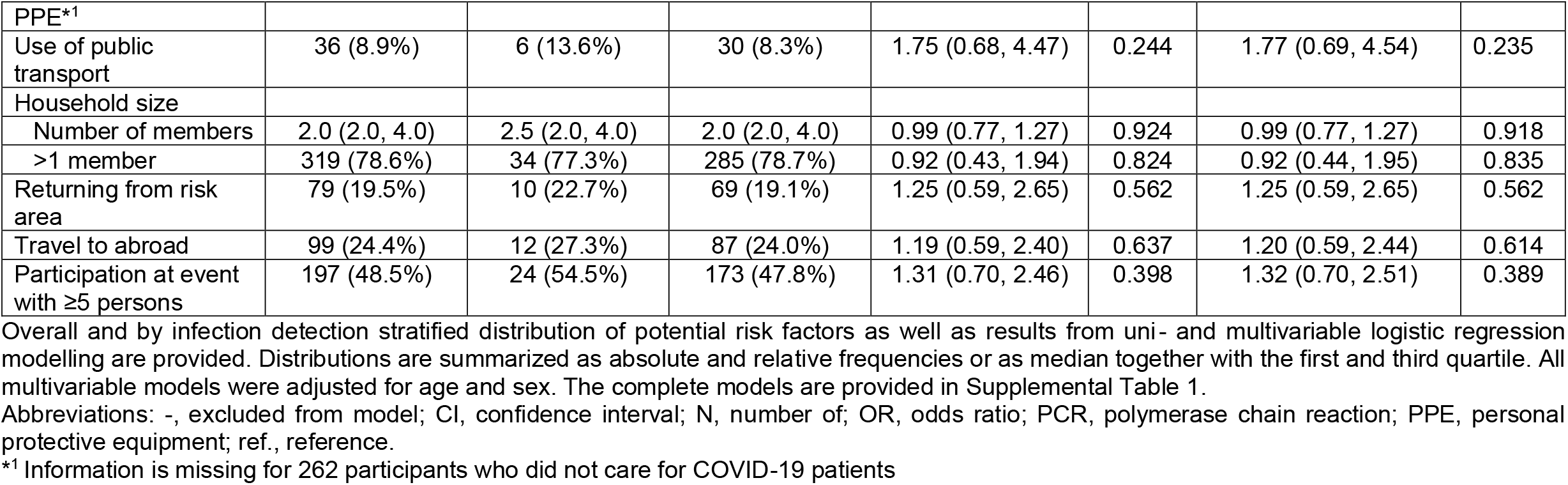
Potential risk factors for a current or past SARS-CoV-2 infection (detected by serology and/or PCR) among hospital staff members

| Variable | Overall<br>(N=406) | Detected infection |  | Univariable analysis |  | Multivariable analysis |  |
| --- | --- | --- | --- | --- | --- | --- | --- |
|  |  | Any (N=44) | None (N=362) | OR (95% CI) | p-value | Adjusted OR (95% CI) | p-value |
| Age, in years | 41.0 (34.0, 49.8) | 43.0 (32.0, 51.0) | 41.0 (34.0, 49.0) | 1.00 (0.97, 1.03) | 0.969 | 1.00 (0.97, 1.03) | 0.962 |
| Male gender | 91 (22.4%) | 11 (25.0%) | 80 (22.1%) | 1.17 (0.57, 2.43) | 0.663 | 1.18 (0.57, 2.43) | 0.663 |
| Profession |  |  |  |  |  |  |  |
| Physician | 66 (16.3%) | 2 (4.5%) | 64 (17.7%) | ref. | 0.107 | ref. | 0.108 |
| Nurse | 125 (30%) | 18 (40.9%) | 107 (29.6%) | 5.38 (1.21, 23.97) | 0.027 | 5.57 (1.24, 25.12) | 0.025 |
| Reception staff | 12 (3.0%) | 1 (2.3%) | 11 (3.0%) | 2.91 (0.24, 34.89) | 0.400 | 3.05 (0.25, 37.65) | 0.348 |
| Administration staff | 132 (32.5%) | 17 (38.6%) | 115 (31.8%) | 4.73 (1.06, 21.13) | 0.042 | 4.92 (1.07, 22.64) | 0.041 |
| Other profession | 71 (17.5%) | 6 (13.6%) | 65 (18.0%) | - | - | - | - |
| COVID-19 risk group according to working place |  |  |  |  |  |  |  |
| High-risk | 76 (18.7%) | 7 (15.9%) | 69 (19.1%) | ref. | 0.643 | ref. | 0.644 |
| Intermediate-risk | 198 (48.8%) | 20 (45.5%) | 178 (49.2%) | 1.11 (0.45, 2.74) | 0.825 | 1.15 (0.46, 2.89) | 0.763 |
| Low-risk | 132 (32.5%) | 17 (38.6%) | 115 (31.8%) | 1.46 (0.58, 3.69) | 0.427 | 1.52 (0.58, 3.98) | 0.397 |
| Reported COVID-19 exposure | 224 (55.2%) | 38 (86.4%) | 186 (51.4%) | 5.99 (2.47, 14.53) | <0.001 | 7.19 (2.86, 18.11) | <0.001 |
| Among them: Place of reported exposure |  |  |  |  |  |  |  |
| Household member | 43 (19.2%) | 16 (42.1%) | 27 (14.5%) | 4.28 (2.00, 9.18) | <0.001 | 4.46 (2.06, 9.65) | <0.001 |
| Friend | 20 (8.9%) | 2 (5.3%) | 18 (9.7%) | 0.52 (0.12, 2.33) | 0.392 | 0.52 (0.11, 2.35) | 0.394 |
| Colleague | 60 (26.8%) | 16 (42.1%) | 44 (23.7%) | 2.35 (1.13, 4.86) | 0.022 | 2.30 (1.10, 4.79) | 0.026 |
| Patient | 151 (67.4%) | 18 (47.4%) | 133 (71.5%) | 0.36 (0.18, 0.73) | 0.005 | 0.36 (0.18, 0.75) | 0.007 |
| Other | 3 (1.3%) | 1 (2.6%) | 2 (1.1%) | 2.49 (0.22, 28.14) | 0.462 | 2.60 (0.22, 30.41) | 0.446 |
| Accident with biological material | 8 (2.0%) | 2 (4.5%) | 6 (1.7%) | 2.83 (0.55, 14.45) | 0.212 | 2.77 (0.54, 14.23) | 0.222 |
| Compliance to wear | 133 (92.4%) | 15 (88.2%) | 118 (92.9%) | 0.57 (0.11, 2.90) | 0.500 | 0.58 (0.11, 2.94) | 0.507 |
| PPE*<sup>1</sup> |  |  |  |  |  |  |  |
| Use of public transport | 36 (8.9%) | 6 (13.6%) | 30 (8.3%) | 1.75 (0.68, 4.47) | 0.244 | 1.77 (0.69, 4.54) | 0.235 |
| Household size |  |  |  |  |  |  |  |
| Number of members | 2.0 (2.0, 4.0) | 2.5 (2.0, 4.0) | 2.0 (2.0, 4.0) | 0.99 (0.77, 1.27) | 0.924 | 0.99 (0.77, 1.27) | 0.918 |
| >1 member | 319 (78.6%) | 34 (77.3%) | 285 (78.7%) | 0.92 (0.43, 1.94) | 0.824 | 0.92 (0.44, 1.95) | 0.835 |
| Returning from risk area | 79 (19.5%) | 10 (22.7%) | 69 (19.1%) | 1.25 (0.59, 2.65) | 0.562 | 1.25 (0.59, 2.65) | 0.562 |
| Travel to abroad | 99 (24.4%) | 12 (27.3%) | 87 (24.0%) | 1.19 (0.59, 2.40) | 0.637 | 1.20 (0.59, 2.44) | 0.614 |
| Participation at event with ≥5 persons | 197 (48.5%) | 24 (54.5%) | 173 (47.8%) | 1.31 (0.70, 2.46) | 0.398 | 1.32 (0.70, 2.51) | 0.389 |

Abbreviations: -, excluded from model; CI, confidence interval; N, number of; OR, odds ratio; PCR, polymerase chain reaction; PPE, personal protective equipment; ref., reference.
\*<sup>1</sup> Information is missing for 262 participants who did not care for COVID-19 patients

### Seroprevalence and prevalence of SARS-CoV-2 infection

At the last study visit, 318 of 406 participants (78.3%) were tested seropositive by Liaison test (295 vaccinated participants and 23 unvaccinated participants), and 88 patients (21.7%) remained seronegative (12 vaccinated participants and 76 unvaccinated participants). Within the 13 months observational period, 44 of 406 participants (10.8%, 95% CI 8.0%-14.3%) had any evidence for a SARS-CoV-2 infection detected by serology and/or PCR. As shown in Table 2, among those 44 participants, 40 participants (90.9%) had at least one positive SARS-CoV-2 IgG antibody test compatible with current or past infection (positive Roche test n=30; positive EDI ELISA n=13; positive Liaison test despite missing vaccination n=26), and 28 participants (63.3%) reported at least one positive PCR test result. According to the self-reported symptoms, nine of the 44 infected participants (20.5%) had an asymptomatic SARS-CoV-2 infection, whereas very mild disease of SARS-CoV-2 related clinical symptoms were reported from two (4.5%), mild disease from eight (18.2%), moderate disease from 14 (31.8%) and severe disease from eleven staff members (25%).

**Table 2.**
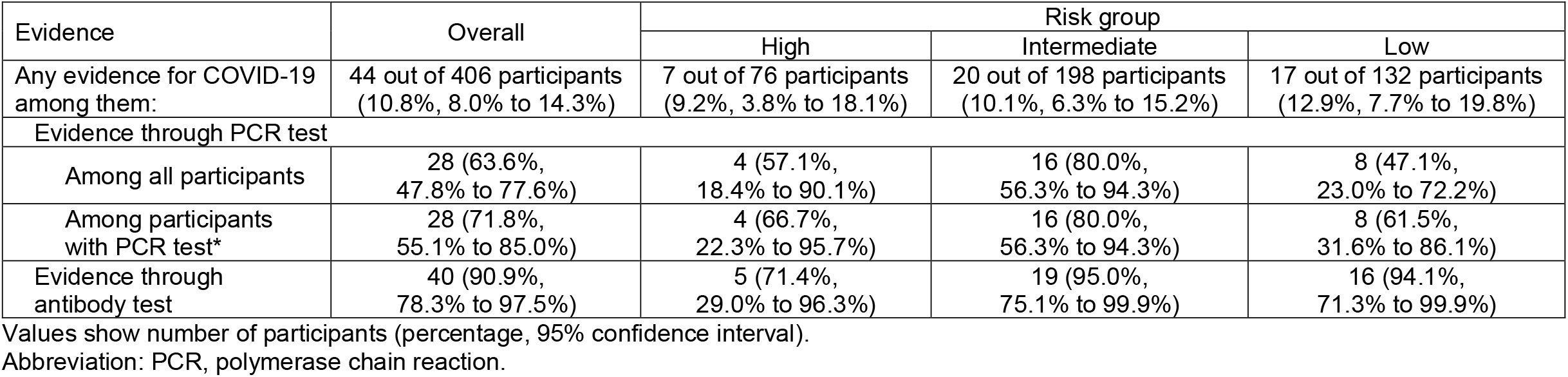
Evidence for a detected COVID-19 infection (PCR and/or antibody test result) among all hospital staff members and stratified for the COVID-19 risk group according to working place

| Evidence | Overall | Risk group |  |  |
| --- | --- | --- | --- | --- |
|  |  | High | Intermediate | Low |
| Any evidence for COVID-19 among them: | 44 out of 406 participants<br>(10.8%, 8.0% to 14.3%) | 7 out of 76 participants<br>(9.2%, 3.8% to 18.1%) | 20 out of 198 participants<br>(10.1%, 6.3% to 15.2%) | 17 out of 132 participants<br>(12.9%, 7.7% to 19.8%) |
| Evidence through PCR test |  |  |  |  |
| Among all participants | 28 (63.6%,<br>47.8% to 77.6%) | 4 (57.1%,<br>18.4% to 90.1%) | 16 (80.0%,<br>56.3% to 94.3%) | 8 (47.1%,<br>23.0% to 72.2%) |
| Among participants with PCR test* | 28 (71.8%,<br>55.1% to 85.0%) | 4 (66.7%,<br>22.3% to 95.7%) | 16 (80.0%,<br>56.3% to 94.3%) | 8 (61.5%,<br>31.6% to 86.1%) |
| Evidence through antibody test | 40 (90.9%,<br>78.3% to 97.5%) | 5 (71.4%,<br>29.0% to 96.3%) | 19 (95.0%,<br>75.1% to 99.9%) | 16 (94.1%,<br>71.3% to 99.9%) |

Values show number of participants (percentage, 95% confidence interval).
Abbreviation: PCR, polymerase chain reaction.

As shown in Figure 1, most positive PCR test results (25/28, 89.2%) were reported during the last six month of the study. SARS-CoV-2 variants of concern (VOCs) alpha, beta, gamma and delta did not emerge among Thuringian surveillance samples earlier than 2021 (alpha variant since January 2021, beta variant since February 2021, gamma and delta variants since April 2021). The molecular surveillance of VOCs and the respected timeline for the State of Thuringia can be assessed at https://charts.mongodb.com/charts-routine-sequencing-sars-c-amykg/public/dashboards/e9453286-1dce-4202-9423-a8459e3962f8.

**Figure 1.**
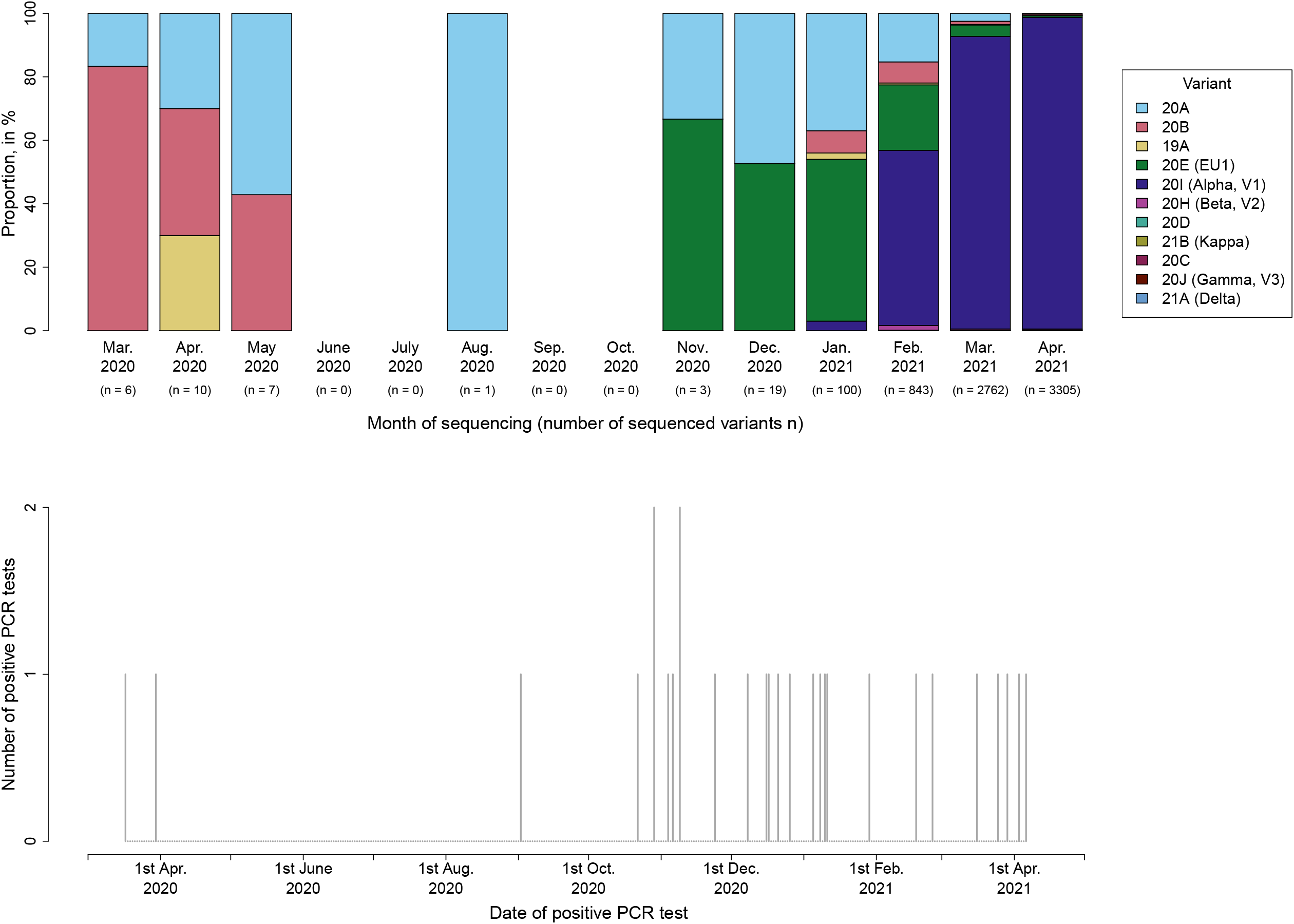
Distribution of SARS-CoV-2 variants from the Thuringian surveillance samples (upper panel) and number and time of reported positive PCR test results among hospital staff members (lower panel) during the period 1^st^ March 2020 to 30^rd^ April 2021. Variants sequenced by the Institute for Infectious Diseases and Infection Control (JUH) are shown. Concerning the data of the SARS-CoV-2 variants, we refer to https://charts.mongodb.com/charts-routine-sequencing-sars-camykg/public/dashboards/e9453286-1dce-4202-9423-a8459e3962f8. Underlying data was assessed at 7^th^ March 2022. Abbreviations: JUH, Jena University Hospital; PCR, polymerase chain reaction.

Two PCR-positive unvaccinated participants did not show any seroconversion. Breakthrough infections after vaccination confirmed by a positive PCR test result were reported in one participant three months after two vaccinations with the COVID-19 mRNA vaccine BNT162b2, in one participant six weeks after one vaccination with the vector based COVID-19 vaccine ChAdOx1-S, and in one participant four months after only one vaccination with BNT162b2.

### Potential risk factors for evidence of any SARS-CoV-2 infection of staff members

As shown in Table 1, we did not find evidence for an association of any current or past SARS-CoV-2 infection (detected by serology and/or PCR) with the demographics, household size, use of public transport to get to work, returning from an inner-German “COVID-19 risk area” as defined by national public health authorities according to the respective incidence, travel to abroad or participation at events with equal to or more than five persons, COVID-19 risk group according to working place, reported accident with biological material or compliance to wear PPE. However, professions associated with an increased risk of experiencing a SARS-CoV-2 infection compared to physicians included nurses (adjusted OR 5.57, 95% CI 1.24-25.12; p=0.025) and administration staff (adjusted OR 4.92, 95% CI 1.07-22.64; p=0.041). Additionally, any reported (occupational and private) COVID-19 exposure (adjusted OR 7.19, 95% CI 2.86-18.11; p<0.001) and particularly close contact to a SARS-CoV-2 positive household member (adjusted OR 4.46, 95% CI 2.06-9.65; p<0.001) and exposure to a SARS-CoV-2 positive colleague (adjusted OR 2.30, 95% CI 1.10-4.79; p=0.026) significantly increased the risk of a SARS-CoV-2 infection among hospital staff. These observations are in line with the results from the additional models for place of exposure, where contact with a household member and with a colleague were both independently associated with a current or past SARS-CoV-2 infection (household member: adjusted OR 5.97, 95% CI 2.07-17.19; p=0.001. Colleague: adjusted OR 3.33, 95% CI 1.36-8.18; p=0.009. Table 3).

**Table 3.** Two additional multivariable logistic regression models for place of exposure

| Variable | Additional model I |  | Additional model II |  |
| --- | --- | --- | --- | --- |
|  | Adjusted OR (95% CI) | p-value | Adjusted OR (95% CI) | p-value |
| Place of reported exposure |  |  |  |  |
| Household member | 5.36 (1.95, 14.77) | 0.001 | 5.97 (2.07, 17.19) | 0.001 |
| Friends | 0.59 (0.09, 3.79) | 0.576 | 0.61 (0.09, 4.15) | 0.615 |
| Colleague | 3.24 (1.33, 7.90) | 0.010 | 3.33 (1.36, 8.18) | 0.009 |
| Patient | 0.91 (0.36, 2.28) | 0.840 | 1.02 (0.38, 2.73) | 0.971 |
| Other | 2.38 (0.10, 55.47) | 0.590 | 2.44 (0.09, 65.87) | 0.596 |
| Age, in years | - | - | 1.02 (0.98, 1.06) | 0.281 |
| Male gender | - | - | 1.09 (0.43, 2.75) | 0.855 |

Abbreviations: -, excluded from model; CI, confidence interval; OR, odds ratio.

## Discussion

The main results of our prospective cohort study among employees at the JUH were the following: (1) The evidence of a past or current SARS-CoV-2 infection detected by serology and/or PCR test results among hospital staff members of JUH tripled from 3.2% during the first corona wave (initial visit ^9^) of the pandemic to 10.8% during the total study period covering the first three corona waves in Germany. This finding is comparable to pooled incidence estimate of SARS-CoV-2 cases of about 12% (95% CI 4%-29%) among HCW reported in a recently published systematic review and meta-analysis with no geographical limitation ^10^. The detected SARS-CoV-2 infection rate in our study was numerically higher compared to the prevalence in the community of the city of Jena. According to the official site of the Robert Koch Institute (https://experience.arcgis.com/experience/478220a4c454480e823b17327b2bf1d4/page/Landkreise/ last accessed at 19^th^ June 2022), the cumulative number of confirmed COVID-19 cases in the city of Jena was 3,902 at 26^th^ April 2021 and 4,382 at 22^nd^ June 2021, corresponding to an infection rate of below 5% of the overall population. However, due to the assessment of seroprevalence and the intense PCR-based HCW screening described, the detection rate at JUH may have been substantially higher compared to the community. (2) We did not identify occupational contact with COVID-19 patients as risk factor for infection. Although the majority of hospital staff members reported direct COVID-19 exposure to a SARS-CoV-2 positive patient (67.4%), there was no evidence for this variable to increase the risk of acquiring an infection, most likely due to a high overall compliance of 92.4% among HCW to wear PPE. HCW caring for COVID-19 patients had a numerically lower infection rate compared to administration staff without any patient care (detected SARS-CoV-2 infection rate: 9.2% among high-risk HCW versus 12.9% among administration staff) and – in line with this observation – patient-related contact to COVID-19 patients was not identified as risk factor in the multivariable analyses. This finding is contradictory to other studies that found a higher absolute risk of seropositivity for HCW with exposure to COVID-19 patients ^3,11,12^. (3) Similar to the first assessment of this study ^9^ and other studies ^3,13^, close contact to a SARS-CoV-2 positive household member was identified as the main private risk factor for a SARS-CoV-2 infection. Additionally, participants with a detected SARS-CoV-2 infection reported more frequently direct exposure to a SARS-CoV-2 positive colleague and were more frequently nurses or administration staff than physicians. The increased infection rate in nurses and administration staff relative to physicians may reflect the impact of medical education on infectious risk assessment and respective risk behaviour including non-patient-related contacts. Even if not addressed in our study, this observation warrants further investigation and may underline the importance of educative measures. Similarly, a recent scoping review that investigated seroprevalence and risk factors of COVID-19 in 9,223 HCW from eleven countries across Africa found that SARS-CoV-2 seropositivity was associated with lower education and working as a nurse/non-clinical HCW ^14^.

This study has the following limitations: Due to the limited number of study visits (two to three per participant within one year) and no mandatory PCR testing among hospital staff, the exact time of SARS-CoV-2 infection detected by serology only could not be determined in 16 hospital staff members and is particularly uncertain in 9 asymptomatic cases. Additionally, underestimation of infection rates could be possible due to waning antibody titres in particular after oligo- or asymptomatic infections ^15,16^.

Hospital staff members may serve as reservoirs, vectors or victims of SARS-CoV-2 cross transmission ^4^. They may not only infect patients they care for but also other HCW, which would cause further reduction of already limited capacity of health services ^3^.

To reduce nosocomial transmissions, the medical executive board of our hospital implemented several specific measures affecting not only the patients but also the hospital staff. For hospital staff, business trips, particularly to travel to abroad, and personal participation on congresses were banned and repeated PCR testing was mandatory when returning from risk areas after holidays. However, these parameters were not associated with an increased risk of SARS-CoV-2 infection in our study. As the own colleagues were identified as the most important source for nosocomial transmissions within the hospital, it was recommended to perform coffee breaks or lunch only with a small number of colleagues with adequate distance and always together with the same colleagues. When mandatory masking was not feasible due to eating, drinking or smoking, speaking should be kept to a minimum.

In conclusion, our results demonstrate that non-patient-related (most-likely non-protected) contacts to SARS-CoV-2 infected household members and colleagues are the main risk factors whereas patient-related contacts (direct contact to COVID-19 patients or body fluids) were not associated with an increased infection risk. Therefore, infection prevention and control strategies should focus more on personal contact between hospital staff members (e.g. using break rooms in small and non-mixed groups only, strict universal masking in team meetings) and should improve risk awareness outside the hospital. The lowest infection rate among physicians compared to nurses and administration employees suggests that medical education may have an impact on risk behaviour also in the non-occupational setting. This underlines the importance of universal masking and educative strategies to decrease the infection risk for hospital employees.

## Data Availability

The datasets used and/or analyzed during the current study are available from the corresponding author on reasonable request.

## Acknowledgements

We thank Steffi Kolanos and Jana Schmidt for excellent technical support.

