## Supplemental file for "Non-patient related SARS-CoV-2 exposure to colleagues and household members impose the highest infection risk for hospital employees with and without patient contact in a German university hospital: follow-up of the prospective Co-HCW Seroprevalence study"

Table S1. Complete multivariable regression models assessing potential risk factors for any evidence of a SARS-CoV-2 infection (detected by serology and/or PCR) among hospital staff members

| Variable | adjusted OR (95% CI) | p-value |
|---|---|---|
| Age, in years | 1.00 (0.97, 1.03) | 0.962 |
| Male gender | 1.18 (0.57, 2.43) | 0.633 |
| Profession*[1] | | |
| Medical doctor | ref. | 0.108 |
| Nurse or care worker | 5.57 (1.24, 25.12) | 0.025 |
| Reception staff | 3.05 (0.25, 37.65) | 0.384 |
| Administration staff | 4.92 (1.07, 22.64) | 0.041 |
| Age, in years | 1.00 (0.96, 1.03) | 0.835 |
| Male gender | 1.13 (0.50, 2.54) | 0.771 |
| COVID-19 risk group according to working place | | |
| High-risk | ref. | 0.644 |
| Intermediate-risk | 1.15 (0.46, 2.89) | 0.763 |
| Low-risk | 1.52 (0.58, 3.98) | 0.397 |
| Age, in years | 1.00 (0.97, 1.03) | 0.826 |
| Male gender | 1.17 (0.56, 2.44) | 0.676 |
| Reported COVID-19 exposure | 7.19 (2.86, 18.11) | <0.001 |
| Age, in years | 1.03 (0.99, 1.06) | 0.122 |
| Male gender | 1.12 (0.53, 2.36) | 0.766 |
| Place of reported exposure: family or partner*[3] | 4.46 (2.06, 9.65) | <0.001 |
| Age, in years | 1.02 (0.99, 1.06) | 0.212 |
| Male gender | 1.02 (0.43, 2.39) | 0.967 |
| Place of reported exposure: friends*[3] | 0.52 (0.11, 2.35) | 0.394 |
| Age, in years | 1.02 (0.98, 1.06) | 0.309 |
| Male gender | 1.04 (0.45, 2.38) | 0.925 |
| Place of reported exposure: colleague*[3] | 2.30 (1.10, 4.79) | 0.026 |
| Age, in years | 1.02 (0.98, 1.05) | 0.402 |
| Male gender | 1.07 (0.47, 2.48) | 0.867 |
| Place of reported exposure: patient*[3] | 0.36 (0.18, 0.75) | 0.007 |
| Age, in years | 1.01 (0.98, 1.05) | 0.528 |
| Male gender | 1.15 (0.50. 2.68) | 0.742 |
| Place of reported exposure: other*[3] | 2.60 (0.22, 30.41) | 0.446 |
| Age, in years | 1.02 (0.98, 1.06) | 0.304 |
| Male gender | 0.95 (0.41, 2.20) | 0.913 |
| Accident with biological material | 2.77 (0.54, 14.23) | 0.222 |
| Age, in years | 1.00 (0.97, 1.03) | 0.945 |
| Male gender | 1.15 (0.55, 2.38) | 0.711 |
| Compliance to wear PPE*[2] | 0.58 (0.11, 2.94) | 0.507 |
| Age, in years | 0.99 (0.94, 1.05) | 0.855 |
| Male gender | 0.96 (0.31, 2.94) | 0.945 |
| Use of public transport | 1.77 (0.64, 4.54) | 0.235 |
| Age, in years | 1.00 (0.97, 1.03) | 0.987 |
| Male gender | 1.20 (0.58, 2.48) | 0.625 |
| Number of household members | 0.99 (0.77, 1.27) | 0.918 |
| Age, in years | 1.00 (0.97, 1.03) | 0.965 |
| Male gender | 1.18 (0.57, 2.43) | 0.661 |
| More than 1 household member | 0.92 (0.44, 1.95) | 0.835 |
| Age, in years | 1.00 (0.97, 1.03) | 0.956 |
| Male gender | 1.17 (0.57, 2.42) | 0.668 |
| Returning from risk area | 1.25 (0.59, 2.65) | 0.562 |
| Age, in years | 1.00 (0.97, 1.03) | 0.942 |

| | | |
|---|---|---|
| Male gender | 1.17 (0.57, 2.43) | 0.664 |
| Travel to abroad | 1.20 (0.59, 2.44) | 0.614 |
| Age, in years | 1.00 (0.97, 1.03) | 0.930 |
| Male gender | 1.19 (0.57, 2.46) | 0.641 |
| Participation at events with ≥5 persons | 1.32 (0.70, 2.51) | 0.389 |
| Age, in years | 1.00 (0.97, 1.03) | 0.832 |
| Male gender | 1.17 (0.56, 2.42) | 0.675 |

[*1] 71 persons with “other profession” were excluded from analysis

[*2] Information is missing for 262 participants

[*3] Among participants with reported place of exposure

Abbreviations: CI, confidence interval; OR, odds ratio; PCR, polymerase chain reaction; PPE, personal protective equipment; ref., reference.
